# Feasibility and efficacy of a digital cognitive behavioural therapy program for insomnia and anxiety for older adults

**DOI:** 10.64898/2025.12.11.25342049

**Authors:** Mathilde Reyt, Jeannick Adoutoro, Florence Borgetto, Caroline Desrosiers, Loïc Barbaux, Kirsten Gong, Josée Savard, Sébastien Grenier, Thien Thanh Dang-Vu

## Abstract

Insomnia and anxiety are highly prevalent and often comorbid in older adults. Although cognitive-behavioural therapy is the first-line treatment for insomnia, few interventions simultaneously address both conditions. Furthermore, access remains limited by provider availability and high costs. To address these gaps, we developed an online CBT program for insomnia and anxiety (eCBT+). This randomized controlled trial aimed to assess the usability and acceptability of the eCBT+ program and evaluate its efficacy in older adults with insomnia.

Eighty older adults with insomnia were randomized to the eCBT+ intervention (*n*=38) or a waitlist (WL) control condition (*n*=42). Platform usability and program acceptability were assessed post-intervention using the System Usability Scale (SUS) questionnaire and the extended Technology Acceptance Model questionnaire. Insomnia and anxiety symptoms evaluated with the Insomnia Severity Index (ISI) and Geriatric Anxiety Inventory (GAI) respectively, along with sleep-diary sleep efficiency, were assessed at baseline and follow-up. Linear mixed models with an intention-to-treat approach assessed the Group*Time interaction.

The platform was considered user-friendly (SUS=69.94%). Perceived ease of use, perceived usefulness, and result demonstrability were the main contributors to acceptability. The eCBT+ group showed reduced ISI and GAI and increased sleep efficiency, from baseline to follow-up, compared to the WL group (*ps*< .001).

The eCBT+ program was user-friendly and its use was acceptable in older adults with insomnia. The program improved sleep efficiency and reduced insomnia and anxiety symptoms, demonstrating the efficacy of our eCBT+ intervention. Web-based tools offer a promising approach to promote sleep and mental health among older adults. (https://www.isrctn.com/ISRCTN15338211)

**KEYPOINTS:**

■ This randomized controlled trial evaluated the feasibility and efficacy of a digital cognitive behavioural therapy program for insomnia and anxiety, delivered through a platform tailored to older adults.
■ The platform was well accepted and considered user-friendly, with acceptability primarily explained by perceived ease of use, perceived usefulness and demonstrability of results.
■ The intervention demonstrated efficacy by improving sleep efficiency and reducing insomnia and anxiety symptoms.
■ This combined digital cognitive behavioural therapy for insomnia and anxiety represents an innovative, clinically relevant and accessible approach for older adults.

## INTRODUCTION

Insomnia disorder is characterized by difficulty initiating or maintaining sleep, accompanied by daytime symptoms [1]. Recent estimates in Canada report that 16 % of adults meet the criteria for an insomnia disorder [2]. This prevalence rises further in older adults due to age-related sleep vulnerability, increased comorbidities and use of medications [3]. In addition, untreated insomnia significantly increases the risk of comorbid mental health disorders [4], such as anxiety [5]. This bidirectional association is reflected in epidemiological data indicating that more than half of adults with anxiety disorder report comorbid insomnia [6], while almost a quarter of individuals with insomnia meet the criteria for an anxiety disorder [7]. The frequent co-occurrence of insomnia and anxiety highlights the need to consider both conditions simultaneously in therapeutic management.

Cognitive-Behavioural Therapy (CBT) is the first-line treatment for insomnia disorder, with demonstrated efficacy in older adults [8] and is also a validated intervention for anxiety [9]. CBT for insomnia targets maladaptive behaviours and dysfunctional beliefs. CBT for anxiety shares cognitive restructuring with which further integrates behavioural activation and stimulus exposure [9]. While previous studies have considered using CBT for a single condition – insomnia or anxiety – in the case of co-occurring disorders [10], there is a lack of data on the combined use of both interventions for individuals experiencing both anxiety and insomnia [11]. This has led to increasing interest in transdiagnostic approaches, which aim to target cognitive-behavioural processes shared across multiple disorders, allowing for integrated treatment of comorbid conditions such as insomnia and anxiety. Indeed, randomized trials have shown that transdiagnostic CBT can significantly reduce insomnia symptoms in university students with emotional disorders [12]. Moreover, emerging neurophysiological frameworks highlight how sleep dysregulation may be a shared mechanistic substrate across anxiety and mood disorders [13].

Face-to-face CBT interventions targeting either insomnia or anxiety have proven effective, improving both sleep and anxiety symptoms [8,9]. However, access to CBT remains limited due to a shortage of trained professionals and its associated costs [14]. To overcome these barriers, an increasing number of digital CBT programs for insomnia have been designed to improve accessibility. Online CBT has been shown to effectively reduce insomnia symptoms, even though its effect may be somewhat lower than those observed in face-to-face or therapist-assisted online interventions [11,15]. Nevertheless, the potential benefits of a combined online intervention for insomnia and anxiety have not yet been established in older adults.

To address this gap, we designed a platform-based CBT program for older adults, addressing both insomnia and anxiety (eCBT+). The first objective of this randomized controlled trial (RCT) was to assess the usability and acceptability of the eCBT+ intervention. Additionally, we aimed to evaluate its efficacy on insomnia and anxiety symptoms in a community sample of older adults randomly assigned to the eCBT+ program or a waitlist control condition (WL). We hypothesized that the eCBT+ intervention would demonstrate high usability and acceptability (i.e., being both easy to use and well received by most participants). Additionally, we expected the eCBT+ group to show greater reductions in insomnia and anxiety symptoms from pre- to post-intervention compared to the waitlist control group.

## METHODS

### Participants and Recruitment

Participants were recruited between October 2021 and February 2023, through the Centre de Recherche de l’Institut Universitaire de Gériatrie de Montréal (CRIUGM) database, social media, newsletters for older adults and clinical referrals. Participants were adults aged 65 years or older, experiencing insomnia symptoms as defined by an Insomnia Severity Index (ISI) score ≥ 8 [16]. They had regular and stable Internet access on a computer, tablet or smartphone to use the online platform. Exclusion criteria included recent hospitalization (within three months), uncorrected visual and/or auditory deficits and conditions preventing study completion, such as suicidal thoughts, neurocognitive disorder or bipolar disorder.

The study protocol was approved by the Research Ethics Committee on Aging – Neuroimaging of the Centre Intégré Universitaire de Santé et de Services Sociaux du Centre-Sud-de-l’Île-de-Montréal and all participants signed an electronic consent form.

### Study design

This study is a RCT (https://doi.org/10.1186/ISRCTN15338211) designed to assess the efficacy of the eCBT+ program. The CONSORT diagram illustrates participant numbers at each stage of the study (Figure 1).

**Figure 1.**
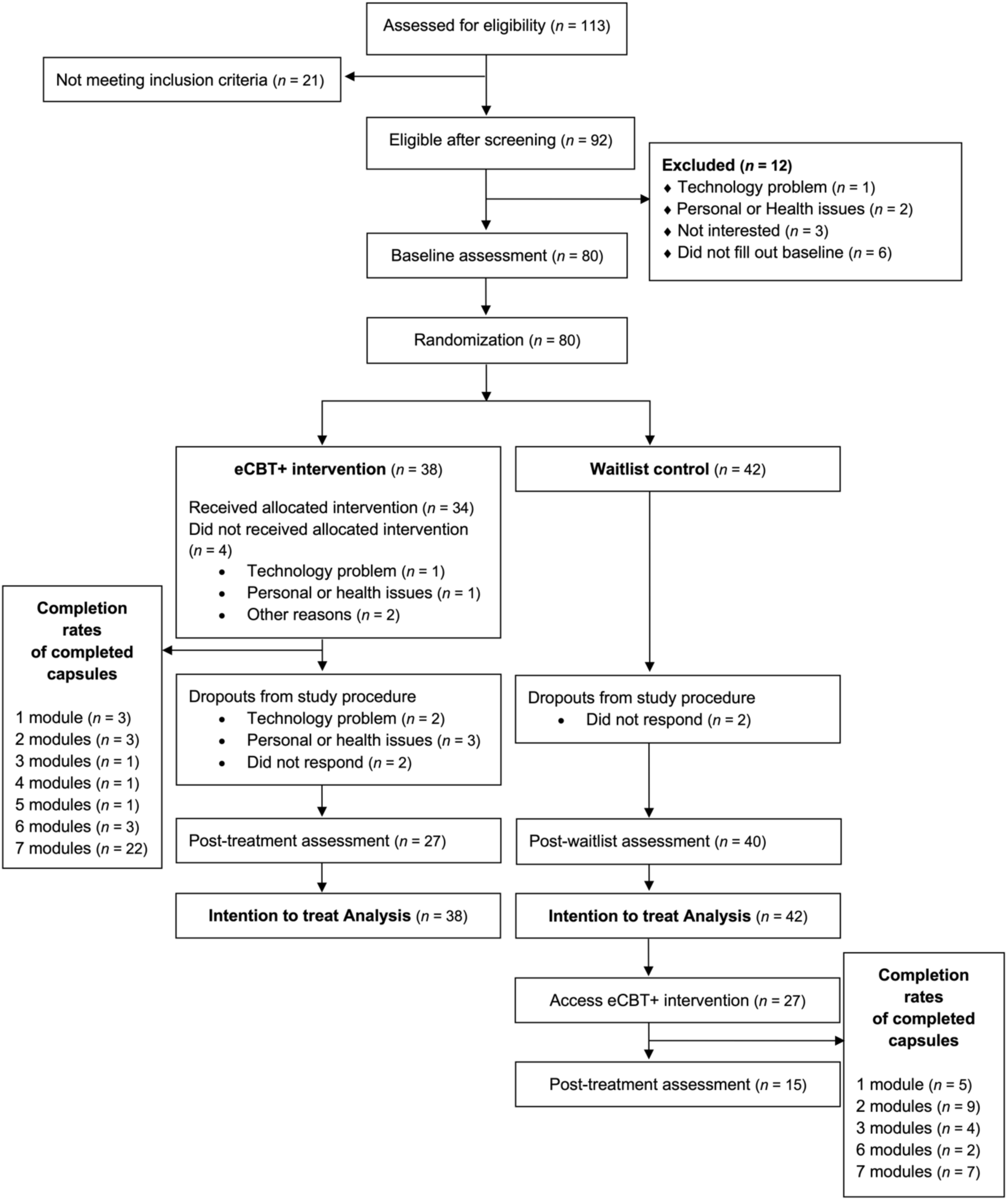
CONSORT diagram depicting participant flow throughout the study

Figure 2 illustrates the study design. Participants’ eligibility was determined through a phone interview covering study explanation and adherence to inclusion/exclusion criteria, alongside collection of demographics (age, height, weight, number of years of education, marital status) and medical data (sleep disorders other than insomnia, use of any medication taken for sleep). Participants with sleep apnoea were not excluded to maintain the ecological validity of the study, as comorbid sleep disorders are common in older adults and previous CBT interventions for insomnia remains effective in the presence of sleep apnoea [17]. At baseline (T1), participants completed online insomnia and anxiety questionnaires and kept a daily sleep diary for two weeks. They were then randomized into either the immediate intervention group (eCBT+ group) or the waitlist control group (WL group). Randomization was stratified based on Geriatric Anxiety Inventory (GAI) score (<7 or ≥7) [18].

**Figure 2.**
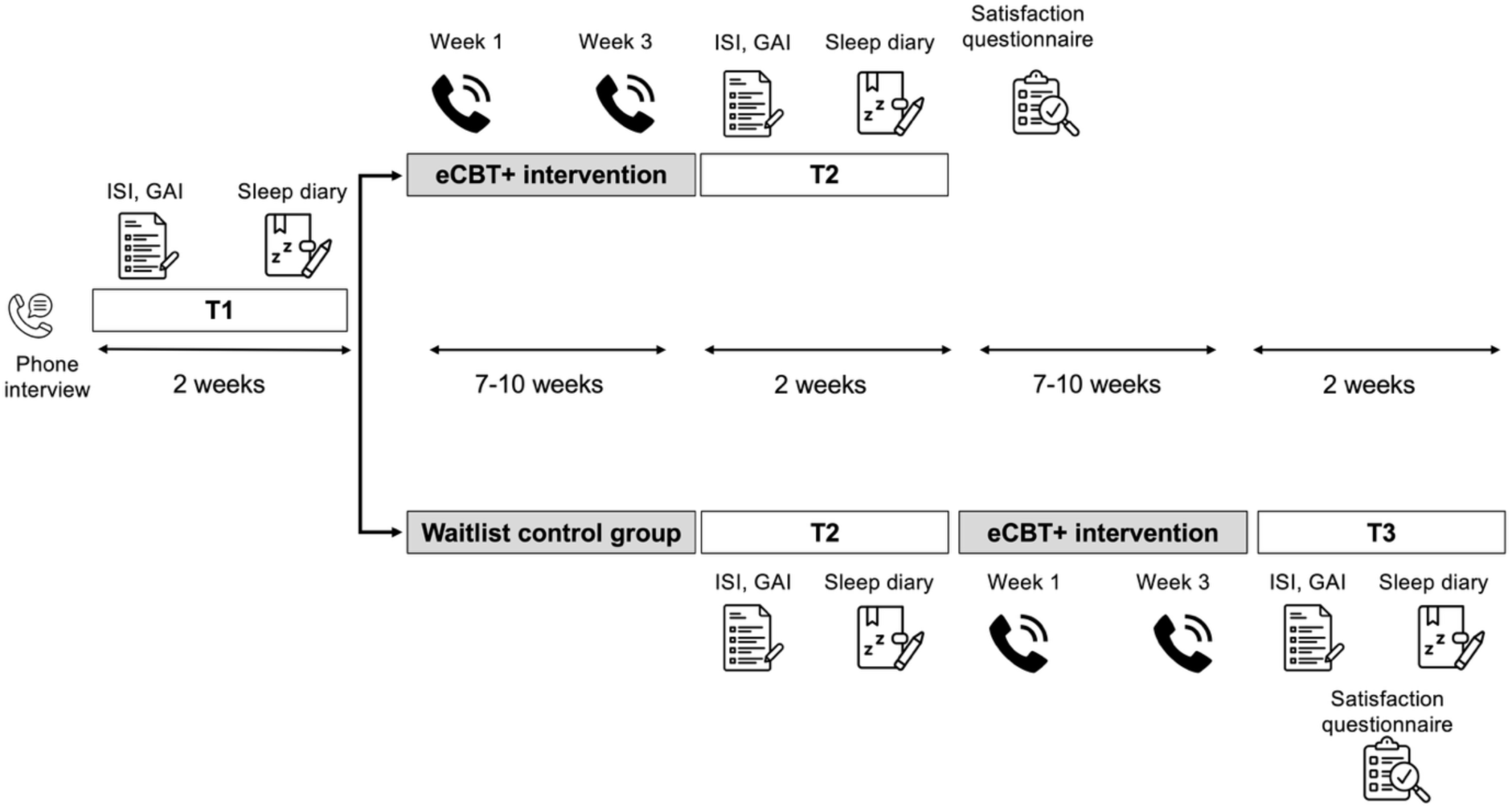
Study protocol. Participants first underwent a phone interview to confirm eligibility. At baseline (T1), they completed online questionnaires assessing insomnia and anxiety (ISI: Insomnia Severity Index; GAI: Geriatric Anxiety Inventory) and maintained a daily sleep diary for two weeks. Participants were then randomized to either the eCBT+ intervention or the waitlist control (WL) group. During the eCBT+ program, participants received two follow-up calls from the research team. After a minimum of seven weeks (T2), both groups repeated the same assessments as at baseline. Subsequently, the WL group was offered the eCBT+ intervention and completed an additional assessment (T3). All participants completed a satisfaction questionnaire upon completion of the eCBT+ program.

After at least 7 weeks, both groups completed a follow-up assessment (T2), similar to the baseline assessment. After completing T2, participants in the WL group were offered the possibility to access the eCBT+ intervention and repeated the assessment (T3).

When registering on the platform, participants were asked to provide information regarding their use of technology. Technology usage was assessed through one single multiple-choice question asking participants to identify the types of technology they commonly used. The available response options were computer, phone, tablet and social networks/social groups. Responses were coded numerically, with one point assigned for each selected option (range from 0 to 4), such that higher scores indicated greater technology use.

Follow-up calls conducted by the research team in weeks 1 and 3 of the eCBT+ program ensured that participants were able to access the logbook activities, clarify questions related to the videos, and discuss their experience in the study. Technical support was provided as needed.

After receiving the eCBT+ intervention, all participants completed a satisfaction questionnaire assessing the platform usability and acceptability.

### Digital CBT program for insomnia and anxiety

Developed at the CRIUGM, the e-SPACE (e-Suite for the Promotion of healthy Aging in the Community) platform is a French-language online platform providing interventions addressing key factors influencing brain health in older adults. The platform is accessible via computer, tablet and smartphone, improving access to brain health interventions to a wide audience. Interventions are divided into four modules (Memory, Communication, Nutrition, and Sleep & Mental Health), with video and interactive content. The present study focuses on the “Sleep and Mental Health” module, developed in collaboration with Dr. Sébastien Grenier and Dr. Josée Savard [19,20]. This represents the eCBT+ intervention and is an online adaptation of CBT for insomnia and anxiety, based on the work of Morin et al. [21,22] and Espie et al. [23] for insomnia and Landreville and colleagues [20] for anxiety. The intervention relies on psychoeducation on insomnia and anxiety, relaxation, cognitive restructuring, problem solving, stimulus control, sleep restriction, sleep hygiene and behavioural activation (see capsules content in Table 1).

**Table 1.** Overview of the eCBT+ intervention capsules.

| **Capsules** | **Title** | **Content** |
| --- | --- | --- |
| 1 | Psychoeducation for anxiety | Definition of anxiety, its causes and the factors influencing its development |
| 2 | Psychoeducation for insomnia | Information on sleep cycles; definition of insomnia, its causes and the factors influencing its development; and guidance on sleep hygiene |
| 3 | Relaxation | Explanation and practical application of two relaxation techniques (progressive muscle relaxation and deep breathing) |
| 4 | Cognitive restructuring | How to identify harmful or unhelpful thoughts and strategies to modify them |
| 5 | Reducing insomnia | Strategies to reduce insomnia, including stimulus control and sleep consolidation techniques |
| 6 | Managing stressors in daily life | Stress management through assertiveness, problem-solving and behavioral activation |
| 7 | Conclusion | Review of key information presented in previous modules, along with the useful therapy resources (e.g., lists of professionals, books on insomnia and anxiety management) |

The eCBT+ intervention was delivered as a self-guided program through seven 30-minute video sessions (capsules) featuring professional voice-over explanations of CBT strategies, animations and interactive elements, quizzes and illustrative patient examples. Participants were instructed to view one capsule per week over seven weeks, with a maximum of ten weeks to account for potential delays in accessing the platform or completing the videos.

### Usability and acceptability of the platform

User experience with the e-SPACE platform was evaluated using a satisfaction questionnaire administered after completion of the eCBT+ program - at T2 for participants in the eCBT+ group (*n* = 25) and at T3 for those in the WL group (*n* = 17). The questionnaire integrated two tools designed to assess the usability of interactive systems, namely the System Usability Scale and the extended model of Technology Acceptance Model.

The System Usability Scale (SUS), developed by Brooke [24], assesses perceived system usability through 10 items covering various aspects of usability (e.g., ease of use, need for support, confidence and consistency of features and functions). Participants rated each item on a 5-point Likert scale (1= strongly disagree, 5= strongly agree), in reference to the e-SPACE platform. Responses to positively worded items were adjusted by subtracting 1 from the raw score, and negatively worded items were adjusted by subtracting the raw score from 5. Adjusted scores for all items were summed and multiplied by 2.5 to yield a total SUS score ranging from 0 to 100, with higher scores indicating greater usability. Cronbach’s alpha values above 0.80 are generally considered indicative of good internal consistency, which was observed in our sample (*α* = 0.88).

The extended model of Technology Acceptance Model (TAM-2) questionnaire [25] is a widely used tool for evaluating technology adoption, assessing behavioural intention to use, perceived usefulness, perceived ease of use, subjective norm, voluntariness, everyday relevance, output quality and results demonstrability. Participants rated each item on a 7-point Likert scale (1= strongly disagree, 7= strongly agree). Positively worded items were scored directly, while negatively worded items were reverse-coded. Total scores ranged from 1 to 7, with higher scores indicating stronger agreement or more favourable perceptions of the system. Overall, the internal consistency of the TAM-2 constructs was satisfactory, with Cronbach’s alpha values ranging from acceptable to excellent for most dimensions (behavioural intention to use = 0.95, perceived usefulness = 0.94, perceived ease of use = 0.88, subjective norm = 0.94, everyday relevance = 0.70, output quality = 0.86 and results demonstrability 0.74). However, the voluntariness construct showed a notably low reliability (α = 0.37).

An additional item assessed participants’ current use of the platform or strategies learned during the intervention, rated on a 5-point frequency scale: 1 = never, 2 = less than once per week, 3 = 1-3 times per week, 4 = 4-7 times per week and 5 = more than 7 times per week.

### Efficacy of the digital CBT program for insomnia and anxiety

Measures assessing the efficacy of the eCBT+ intervention were collected at T1 and T2, and at T3 for participants in the WL group.

Over a 14-day period, participants completed a daily sleep diary [26] that included bedtime, lights-out time, sleep onset latency, wake after sleep onset, wake-up time and out-of-bed time. Time in bed (TIB) and total sleep time (TST) were calculated from these entries to derive sleep efficiency (SE = TST/TIBx100).

The Insomnia Severity Index (ISI) [16] is a 7-item self-report questionnaire assessing the nature, severity and impact of insomnia symptoms. Items address difficulty falling asleep, difficulty staying asleep, problems with early morning awakenings, satisfaction with current sleep pattern, interference with daily functioning and distress caused by sleep difficulties. Each item is rated on a 5-point Likert scale (0= no problem, 4= very severe problem), yielding a total score from 0 to 28. Higher scores indicate greater insomnia severity. The ISI demonstrated satisfactory internal consistency, as indicated by Cronbach’s alpha values of 0.82 at T1, 0.87 at T2 and 0.91 at T3.

In addition to assessing symptom severity, the ISI also allows the classification of clinically meaningful improvements. Participants were classified as responders if they showed a reduction of at least 7 points on the ISI from baseline, reflecting a clinically meaningful improvement in insomnia symptoms. Remitters were defined as participants whose post-treatment ISI score fell below the established threshold for insomnia (ISI ≤ 7), indicating the resolution of clinically significant symptoms.

The Geriatric Anxiety Inventory (GAI) [18] is a 20-item self-report questionnaire developed to assess anxiety symptoms in older adults. Each item is answered with a dichotomous “agree” or “disagree” response and scored 1 for agreement with an anxiety-related statement and 0 for disagreement, yielding a total score ranging from 0 to 20. Higher scores indicate greater anxiety severity. Internal consistency of the GAI was found to be good, as indicated by Cronbach’s alpha values (*α* = 0.86 at T1, *α* = 0.89 at T2 and *α* = 0.88 at T3).

### Statistical analyses

For the usability and acceptability of the platform, data from both groups were analysed together using descriptive statistics only. Means and standard deviations were computed for all questionnaire scores. No inferential analyses were performed, as the aim was to provide an overview of usability and acceptance of the e-SPACE platform. To examine the effects of the eCBT+ intervention on sleep efficiency, ISI and GAI scores, linear mixed-effects models were conducted for each outcome variable separately. Each model included Group (eCBT+ vs. WL), Time (Pre vs. Post intervention), and their interaction (Group × Time) as fixed effects, with age and sex as covariates. A random intercept for subject was included to account for repeated measures within individuals. Significance of fixed effects was tested using Type IIII analyses of variance (ANOVA). When significant Group x Time interactions were observed, post-hoc comparisons were conducted using estimated marginal means (EMMs). Between-group Cohen’s *d* values were computed from the differences between EMMs of the groups, using the model’s residual standard deviation as the standardizer. Within-group Cohen’s *d* values were calculated for pre-to-post changes within each group. For all Cohen’s *d* effect sizes, a positive value indicates an improvement in the outcome. Effect sizes were interpreted according to conventional thresholds: 0.2 = small, 0.5 = medium, and 0.8 = large. Confidence intervals were reported alongside Cohen’s *d* to provide an estimate of precision. Data were analysed with an intention-to-treat approach, whereby all randomized participants were included in the analyses according to their assigned group. Graphical diagnostics and Shapiro-Wilk test were used to check the normality of the distribution of each outcome variable. Levene’s test was used to examine homogeneity of variance. All analyses were performed using R version 4.4.1 [27].

To further confirm the effect sizes of the eCBT+ intervention, we combined T2 data from participants initially receiving eCBT+ with T3 data from participants initially assigned to the waitlist who subsequently received the intervention. Linear mixed-effects models were used for each outcome, including Time as a fixed effect, and age and sex as covariates. A random intercept for each subject was specified to account for repeated measures within individuals. Type III analysis of variance (ANOVA) was performed on each model to evaluate the significance of main effect. This approach enabled the assessment of changes over time while adjusting for individual differences and covariates. Effect sizes were estimated using within Cohen’s *d* and were reported alongside their respective confidence intervals to provide an estimate of precision.

## RESULTS

### Socio-demographic

As shown in the CONSORT (Figure 1), a total of 113 participants were screened via telephone. Of these, 21 individuals were excluded for not meeting the inclusion criteria. Of the 92 eligible participants, 1 experienced technical problems, 2 withdrew due to personal or health issues, 3 were not interested in participating and 6 did not complete the baseline assessment and were therefore not included in the study.

A total of 80 participants were then randomized: 38 in the eCBT+ group and 42 in the WL group. In the eCBT+ group, 4 participants did not start the intervention: 1 withdrew due to personal or health reasons, 1 due to difficulties registering on the platform and 2 for other reasons. Among those who started the intervention, 2 participants discontinued because of technology or platform problems and 3 withdrew for personal or health-related reasons. Overall, 29 out of the 38 participants randomized to the eCBT+ group completed the intervention, corresponding to a retention rate of 76.32%. Furthermore, two participants from the eCBT+ group and two from the WL group did not complete the post-treatment or post-waitlist assessment, respectively. Participants who did not complete the study did not differ significantly from completers in terms of demographic characteristics, baseline ISI, GAI or sleep efficiency (see Table S1).

Descriptive statistics for demographic characteristics are presented in Table 2. The randomized sample consisted of 80 participants, predominantly women (76.25%), with a mean age of 71.86 ± 5.33 years (range: 65 and 84 years). A large proportion had completed university education (83.75%). More than half were married (53.75%) and reported taking medication to help them sleep (60%). Sleep apnoea was self-reported by 17.5% of participants. More than half of the participants (55%) reported using at least three different devices, indicating that the sample was generally comfortable with technology. No group difference was found for demographical variables (all *p*_s_> 0.05, see Table 2), with a trend for higher sleep apnoea in the waitlist group.

**Table 2.**
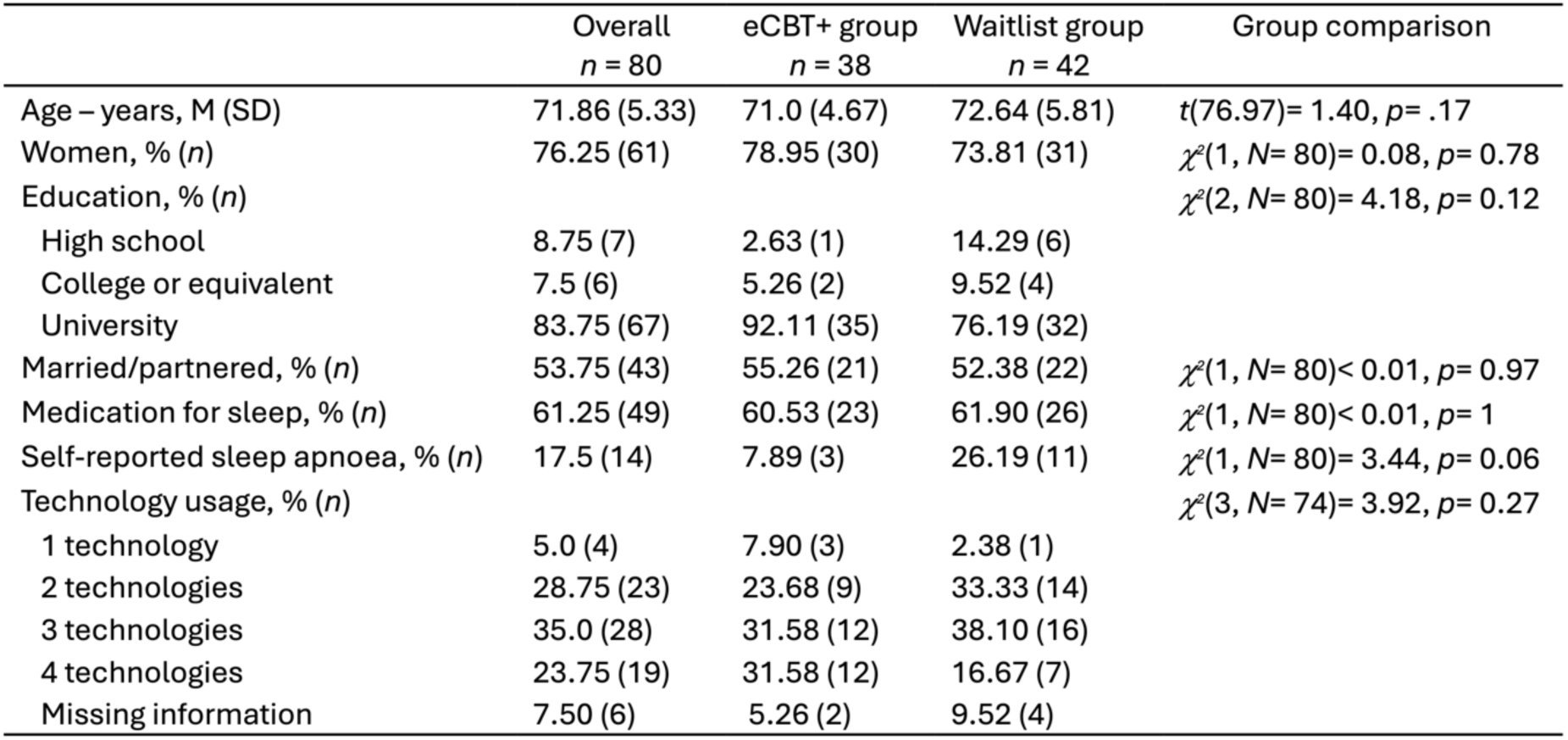
Sociodemographic and clinical characteristics of participants.

|  | Overall<br>*n* = 80 | eCBT+ group<br>*n* = 38 | Waitlist group<br>*n* = 42 | Group comparison |
| --- | --- | --- | --- | --- |
| Age – years, M (SD) | 71.86 (5.33) | 71.0 (4.67) | 72.64 (5.81) | $t(76.97) = 1.40, p = .17$ |
| Women, % (*n*) | 76.25 (61) | 78.95 (30) | 73.81 (31) | $\chi^2(1, N = 80) = 0.08, p = 0.78$ |
| Education, % (*n*) | | | | $\chi^2(2, N = 80) = 4.18, p = 0.12$ |
| High school | 8.75 (7) | 2.63 (1) | 14.29 (6) |  |
| College or equivalent | 7.5 (6) | 5.26 (2) | 9.52 (4) |  |
| University | 83.75 (67) | 92.11 (35) | 76.19 (32) |  |
| Married/partnered, % (*n*) | 53.75 (43) | 55.26 (21) | 52.38 (22) | $\chi^2(1, N = 80) < 0.01, p = 0.97$ |
| Medication for sleep, % (*n*) | 61.25 (49) | 60.53 (23) | 61.90 (26) | $\chi^2(1, N = 80) < 0.01, p = 1$ |
| Self-reported sleep apnoea, % (*n*) | 17.5 (14) | 7.89 (3) | 26.19 (11) | $\chi^2(1, N = 80) = 3.44, p = 0.06$ |
| Technology usage, % (*n*) | | | | $\chi^2(3, N = 74) = 3.92, p = 0.27$ |
| 1 technology | 5.0 (4) | 7.90 (3) | 2.38 (1) |  |
| 2 technologies | 28.75 (23) | 23.68 (9) | 33.33 (14) |  |
| 3 technologies | 35.0 (28) | 31.58 (12) | 38.10 (16) |  |
| 4 technologies | 23.75 (19) | 31.58 (12) | 16.67 (7) |  |
| Missing information | 7.50 (6) | 5.26 (2) | 9.52 (4) |  |

### Usability and acceptability of the platform for the digital CBT program for insomnia and anxiety

The SUS yielded a mean score of 69.94% (± 19.3) for the overall sample (*n* = 42) after completion of the eCBT+ program in both the eCBT+ group (*n* = 25) and the WL group (*n* = 17). According to Bangor et al. [28], SUS scores can be interpreted using the acceptability scale: scores below 50 are considered “not acceptable”, scores between 50 and 69 are “marginal” and scores of 70 and above are “acceptable”. On this scale, the e-SPACE platform falls within the high end of the “marginal” range, close to the threshold for “acceptable” usability.

The TAM-2 analysis (Figure 3) revealed a high behavioural intention to use the system (5.96 ± 1.16 on a 7-point Likert scale), suggesting strong willingness among participants to adopt the platform. Perceived ease of use was also rated positively (5.40 ± 1.33), indicating that users generally found the system intuitive. Perceived usefulness (5.04 ± 1.27) and everyday relevance (4.95 ± 1.46) received favourable evaluations, reflecting the perception that the platform supports meaningful and contextually relevant tasks. Results demonstrability scored similarly high (5.08 ± 1.05), highlighting that users felt the benefits of the system were evident and measurable. In contrast, subjective norm (4.45 ± 1.29), voluntariness (4.37 ± 0.79), and output quality (4.37 ± 1.59) were comparatively lower, suggesting that social influence, perceived autonomy, and satisfaction with the system’s outputs contributed less to acceptance. Overall, the strongest factors of acceptability appeared to be perceived ease of use, perceived usefulness, and the demonstrability of results. 40.5% of participants reported using the e-SPACE platform or its taught strategies between 4-7 times per week, with an additional 14.3% indicating usage more than seven times per week. Less participants reported using it 1-3 times per week (19.0%), less than once per week (14.3%), or never (11.9%). These findings highlight that most participants integrated e-SPACE into their routines on a frequent—often near-daily—basis.

**Figure 3.**
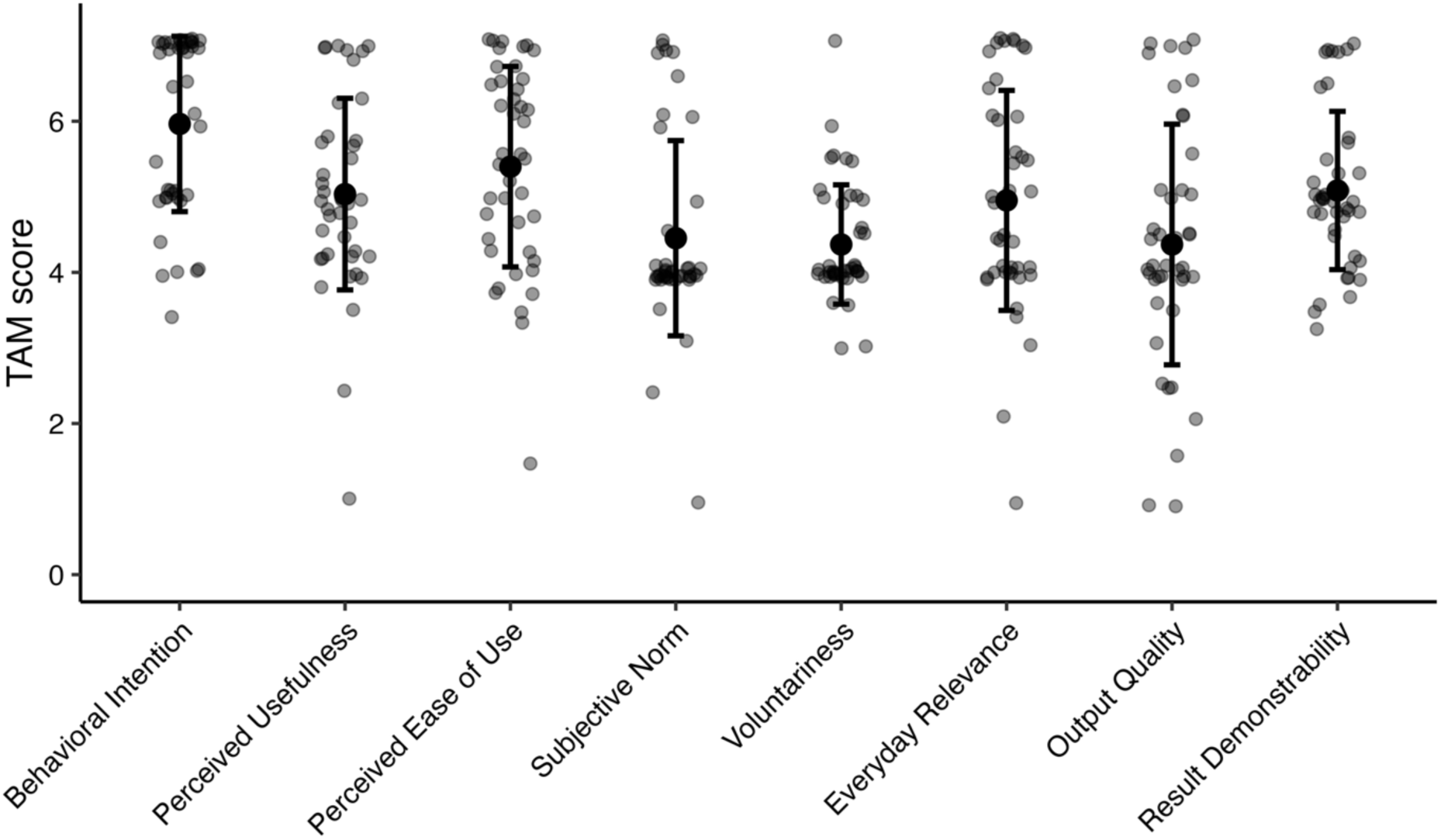
Technology Acceptance Model (TAM-2) scores for each domain, showing the mean ± standard deviation and individual participant scores (grey points). Scores are based on a 7-point Likert scale, with higher values indicating stronger agreement.

### Efficacy of the digital CBT program for insomnia and anxiety

The linear mixed-effects models revealed a significant Group*Time interaction for sleep efficiency (*F*(1, 63.91) = 30.83, *p*< .001, Table 3). The eCBT+ intervention led to a significant improvement in sleep efficiency compared to the WL group from pre- to post-intervention (Figure 4A, *t*(75.7) = 2.76, *p*< .01, Cohen’s *d_between_* = 1.24, 95% CI [0.33, 2.15]).

**Figure 4.**
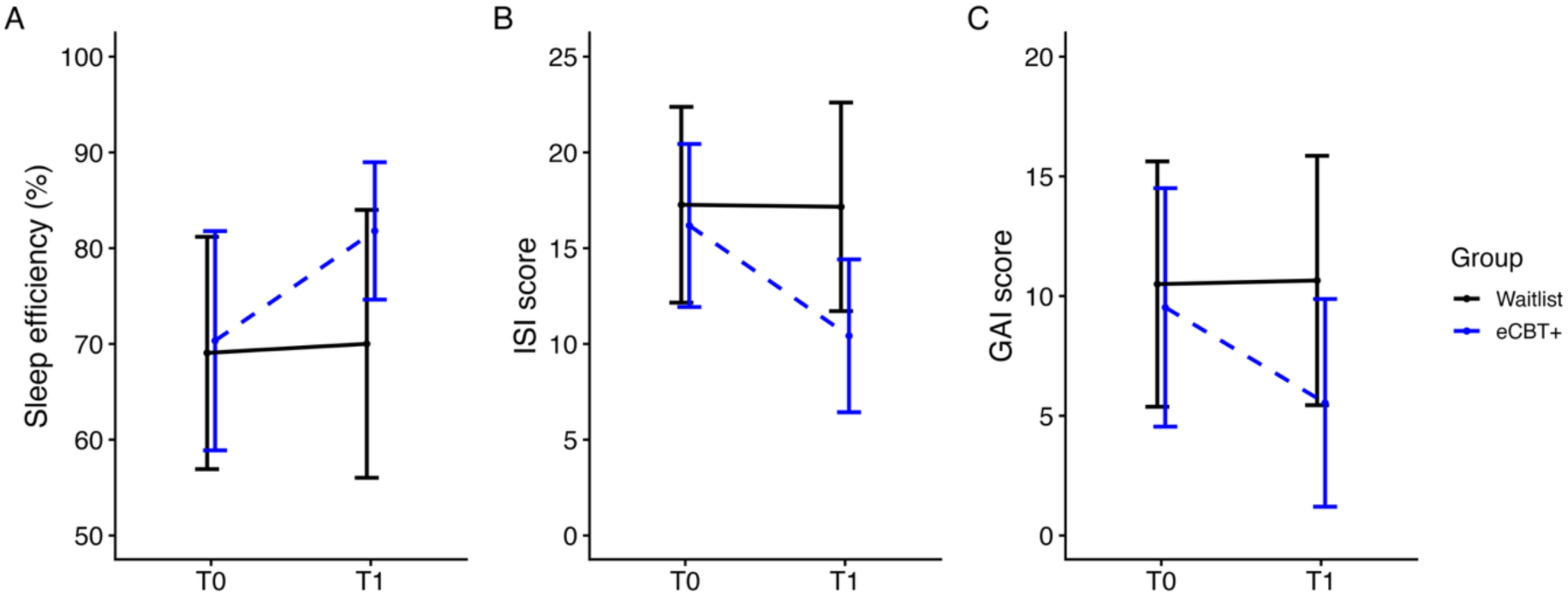
Effects of the eCBT+ program on sleep-diary sleep efficiency (A), insomnia severity (ISI) (B), and anxiety symptoms (GAI) (C). The eCBT+ group is shown in blue with a dashed line, while the waitlist control group is shown in black with a solid line.

**Table 3.** Effects of the eCBT+ program on sleep-diary sleep efficiency, insomnia severity (ISI), and anxiety symptoms (GAI). Values are presented as mean with standard deviation (SD) for each group and timepoint. Effects sizes are reported with 95% confidence intervals (CI), as well as linear mixed model outputs for the Time x Group interaction, including β, standard error (SE) and p-value.

| Outcome/Timepoint | eCBT+ group |  |  | Waitlist group |  |  | Time x Group |  |  |  |
| --- | --- | --- | --- | --- | --- | --- | --- | --- | --- | --- |
| | n | Mean (SD) | Cohen's $d_{within}$ [95% CI] | n | Mean (SD) | Cohen's $d_{within}$ [95% CI] | $\beta$ | SE | p value | Cohen's $d_{between}$ [95% CI] |
| Sleep efficiency (%) |  |  |  |  |  |  |  |  |  |  |
| T0 | 37 | 70.34 (11.45) | 1.20 [0.68, 1.71] | 42 | 69.06 (12.13) | 0.03 [-0.35, 0.30] | -11.24 | 2.02 | $p < .001$ | 1.24 [0.33, 2.15] |
| T1 | 25 | 81.8 (7.17) |  | 37 | 70.01 (13.98) |  |  |  |  |  |
| Insomnia severity |  |  |  |  |  |  |  |  |  |  |
| T0 | 38 | 16.18 (4.25) | 1.0 [0.52, 1.47] | 41 | 17.27 (5.11) | 0.04 [-0.28, 0.35] | 5.11 | 1.15 | $p < .001$ | 1.18 [0.56, 1.79] |
| T1 | 26 | 10.42 (3.99) |  | 38 | 17.16 (5.45) |  |  |  |  |  |
| Anxiety severity |  |  |  |  |  |  |  |  |  |  |
| T0 | 38 | 9.53 (4.98) | 0.74 [0.30, 1.17] | 40 | 10.5 (5.12) | 0.05 [-0.27, 0.37] | 3.48 | 0.95 | $p < .001$ | 1.17 [0.38, 1.96] |
| T1 | 26 | 5.54 (4.34) |  | 37 | 10.65 (5.21) |  |  |  |  |  |

Similarly, a significant Group*Time interaction was observed for the ISI questionnaire (*F*(1, 65.84) = 19.75, *p*< .001, Table 3). From pre- to post-intervention, the eCBT+ group showed significantly lower ISI scores compared to the WL group (Figure 4B, *t*(75.3) = −3.93, *p*< .001, Cohen’s *d_between_*= 1.18, 95% CI [0.56, 1.79]).

Clinically meaningful response was observed in 13 of 26 participants in the eCBT+ group, compared with 2 of 38 in the WL group. This difference was statistically significant (*χ*²(1, N = 64) = 14.82, *p*< .001), reflecting a higher response rate in the eCBT+ group. Among participants in the eCBT+ group, 5 of 26 met criteria for remission, while 3 of 38 did so in the WL group. Remission rates were therefore comparable across groups, with no significant difference between groups (*χ*²(1, N = 64) = 0.93, *p* = 0.34).

Analysis also revealed a significant Group*Time interaction on the GAI scores (*F*(1, 65.84) = 13.46, *p*< .001, Table 3), with the eCBT+ group showing a decrease in anxiety scores from pre- to post-intervention compared to the WL group (Figure 4C, *t*(74.6) = −2.99, *p*< .01, Cohen’s *d_between_*= 1.17, 95% CI [0.38, 1.96]).

As we observed a trend toward a between-group difference in self-reported sleep apnoea, we performed additional analyses on efficacy after excluding participants with self-reported sleep apnoea in each group. Similar results were obtained, with large effect sizes also observed (see Table S2).

To further confirm the effects of the eCBT+ intervention, we combined T2 data from participants initially receiving eCBT+ with T3 data from participants initially assigned to the waitlist who subsequently received the intervention. Analyses (Table S3) revealed a significant effect of Time, with increased sleep efficiency (*F*(1, 41.34) = 38.11, *p*< .001, Cohen’s *d_within_* = 1.41, 95% CI [0.91, 1.91]), and reductions in ISI (*F*(1, 43.26) = 33.82, *p*< .001, Cohen’s *d_within_*= 1.25, 95% CI [0.78, 1.72]) and GAI (*F*(1, 41.96)= 21.20, *p*< .001, Cohen’s *d_within_* = 1.03, 95% CI [0.56, 1.51]) scores after the eCBT+ intervention.

## DISCUSSION

In a sample of older adults (mainly women) with mild to moderate insomnia and comorbid anxiety, we demonstrated that the first French-language online CBT program targeting both insomnia and anxiety (eCBT+) was user-friendly and well accepted. This intervention was also effective in improving sleep efficiency and reducing insomnia and anxiety symptoms compared to a waitlist group receiving no intervention.

For comparison, we summarized the data from other RCTs reporting the effects of digital CBT interventions for insomnia on sleep efficiency, ISI or anxiety symptoms (Supplemental Table S4). For each study, retention rate, pre-to-post changes and between-group effect sizes (Cohen’s *d*) are reported, to facilitate comparison with the present findings.

### Usability and acceptability of the digital CBT program for insomnia and anxiety

Our results show that the web-based eCBT+ program reached a sufficient level of usability (∼70%), supported by participants’ perceived ease of use, perceived usefulness and demonstrability of results. Participants highlighted several positive aspects of the platform, noting its simplicity, ease of learning, the confidence it generated while using it and the good integration of its various functions, which encouraged frequent use. Technology usage levels did not influence therapy completion, as there were no significant differences in the number of technologies commonly used between participants who completed the program and those who withdrew.

Although there is a growing body of research on digital CBT for insomnia [11], we identified only three studies that specifically assessed usability using the SUS questionnaire, reporting score of 80.42 [29], 80.7 [30] and 85.74 [31]. Compared with these studies, our eCBT+ program achieved a slightly lower but still acceptable score, which may be explained by the fact that the questionnaire assessed the usability of the entire platform. Participants navigated through several types of content on the eSPACE platform – videos, questionnaires and video-related activities – which may have introduced additional navigation difficulties. The relatively modest influence of social factors on intervention acceptance may be explained by the therapeutic nature of the program. Psychological treatments are typically perceived as personal and sensitive, and individuals may neither expect nor desire social endorsement for engaging in such interventions. In this context, digital healthcare may offer the advantage of allowing greater privacy, thereby reducing concerns about social judgment [32].

Factors positively influencing the acceptability of our eCBT+ intervention were participants’ behavioural intention to use it, its perceived ease of use, its perceived usefulness, and demonstrability of results. Ease of use and perceived usefulness are recognized as key drivers of technology adoption, particularly among older adults using health information technologies [33] and the present study represents a novel application of the TAM-2 to assess the acceptability of insomnia and anxiety therapies. Consistent with prior findings in digital healthcare, both perceived usefulness and ease of use positively influenced attitudes toward use in older adults [34], while results demonstrability – participants observing the effects of eCBT+ – also promoted platform adoption.

The retention rate in the eCBT+ group was 76.32%, which falls within the range reported in previous digital CBT trials for insomnia (43-100%) and is comparable to the average retention rate typically observed across studies (around 80-85%, see Table S4). No significant differences were observed between participants who dropped out and those who completed the intervention in terms of demographic characteristics or baseline sleep efficiency, ISI and GAI scores. Dropouts were attributable to both health-related and technical factors, with 2 participants withdrawing due to difficulties with registration and navigation on the platform. A few participants expressed feeling discouraged by the lack of noticeable improvement, particularly if they were already familiar with the therapy content, which may have reduced engagement. These findings mirror those reported by Simon and colleagues [35], who similarly found that none of the examined baseline variables significantly predicted dropout in their digital CBT program for insomnia, with most participants citing distractions from daily life (65.4%), technical difficulties (28.6%) and perceptions that the intervention was not helpful for improving sleep-related symptoms (45.5%) as main reasons for discontinuation.

Overall, these results suggest that our eCBT+ intervention was a well-accepted digital intervention for insomnia, even among participants who may encounter typical obstacles to engagement.

### Efficacy of the digital CBT program for insomnia and anxiety

The eCBT+ intervention proved effective in improving sleep and anxiety in participants receiving the eCBT+ program, compared to a control group, with large effect sizes observed across outcomes.

Following the intervention, average sleep efficiency in the eCBT+ group exceeded 80%, while it remained stable at 70% in the waitlist group. The average 11.46% improvement in sleep efficiency observed in our sample is consistent with effects reported in previous digital CBT trials for insomnia (see Table S4). The between-group effect size for sleep efficiency in our study is among the highest reported, highlighting the substantial impact of our intervention. The eCBT+ intervention also improved insomnia symptoms, with ISI scores decreasing by 5.76 points. The effect size observed for ISI is within the upper-middle range of effect sizes reported in previous trials (see Table S4), indicating a large and clinically meaningful impact. Notably, this effect size is similar to the one observed in the only previous RCT evaluating digital CBT for insomnia focusing on older adults [36].

The eCBT+ intervention also reduced anxiety symptoms, decreasing GAI scores by 3.4 points on average. Previous RCTs of digital CBT programs for insomnia have evaluated anxiety using different measures, none matching the questionnaire employed in our study (see Table S4). In this context, the between-group effect size for anxiety symptoms in our sample is among the highest reported, suggesting a particularly robust impact. Although previous digital CBT trials for insomnia have documented beneficial effects on anxiety, the particularly large effect observed in our sample indicates that the joint improvement of insomnia and anxiety represents a key strength of our program.

Our results validate the efficacy of the eCBT+ intervention for treating insomnia and anxiety. Participant feedback may help guide future adaptations of this intervention; for instance, one participant suggested allowing more time to review the material. The seven-week duration of our program falls within the typical range, as most RCTs evaluating digital CBT interventions for insomnia have implemented six-week programs (see Table S4), although programs with 10 sessions have demonstrated the largest effect sizes [37,38,39,40], in line with moderator analyses showing that longer treatment duration is associated with larger effect sizes [41]. This suggests that extending the number of sessions may further enhance the efficacy of digital CBT interventions for insomnia and should be considered in future adaptations. Occasional email, phone or in-app notifications may also enhance adherence, a strategy already used in most online CBT programs for insomnia (e.g., NUKKUAA, SHUTi, Sleepio, Somnio, Somnovia and Somzz). Finally, enhancing participant follow-up by involving trained professionals could help address questions about therapeutic content via discussion forum, chat, or email. Supporting findings from a meta-analysis of internet-based CBT studies for insomnia showed that greater personal clinical support is associated with larger effect sizes [41]. Optimizing therapeutic approaches for older adults also requires consideration of factors such as cognitive decline, multimorbidity, and internalized ageism, which can affect efficacy [42]. Considering the high prevalence of depression among older adults with anxiety and/or sleep disturbances, future studies could also assess comorbid depressive symptoms. One participant in our study, for instance, suggested providing session summaries to help retain key points.

### Strengths and limitations

This study presents several key strengths. Our eCBT+ intervention uniquely combines components and strategies targeting both insomnia and anxiety, addressing their frequent comorbidity and reciprocal influence, which is particularly relevant in older adults. Our platform and intervention were carefully tailored for this population, with adjustable features, such as text size and contrast, and scenario-based illustrations relevant to the daily lives of older adults. Furthermore, the RCT design strengthens the validity of our findings, allowing for accurate estimation of the eCBT+ intervention’s effects and effect sizes while controlling for confounding factors.

In our sample, over 83.75% of participants had a high education level, substantially higher than the proportion of Canadians aged 65 and over with post-secondary education in 2021, which was less than 50% [43]. This high level of education may have influenced not only participants’ evaluation of the online intervention’s usability and acceptability but also their general technology use. Indeed, a Canadian study on technology use for social interaction among older adults found that higher education was associated with greater awareness and uptake of technology, suggesting that more educated participants may be more comfortable and confident with digital tools [44]. Moreover, educational level may also shape treatment response to digital CBT for insomnia: in a recent digital CBT trial for insomnia, individuals with lower education showed markedly lower odds of remission [45]. As a result, this characteristic may limit the generalizability of our findings to older adults with lower educational level or less experience with technology. Another limitation concerns the French-only availability of the intervention. Developing an English version would expand its accessibility and allow a broader range of older adults, both within Canada and internationally, to benefit from the eCBT+ program.

## Conclusion

Our findings demonstrate that the eCBT+ program is not only usable and acceptable but also highly effective, yielding large improvements in insomnia severity, anxiety and sleep efficiency. These robust outcomes underscore both the clinical significance and practical utility of the intervention, establishing the eCBT+ program as a highly promising and accessible approach for addressing insomnia and anxiety in older adults.

Future studies will confirm these effects in broader samples of older adults with insomnia. Such studies may examine effects of the eCBT+ intervention on daytime functioning, and particularly cognitive performance, as key aspects of insomnia-related quality of life. Additionally, follow-up evaluations are needed to confirm the persistence of eCBT+ effects over the long term.

## AUTHORS CONTRIBUTIONS

MR, FB, JS, SB, and TDV conceptualized the study. JA, FB, CD, LB, and KG were responsible for recruitment and data collection. Project coordination was managed by JA and FB. Data management was performed by JA, FB, and MR. MR conducted the statistical analyses. MR, FB, and TDV interpreted the results. The original draft of the manuscript was written by MR, FB, and TDV. All authors contributed to critically reviewing and editing the manuscript and approved the final version.

## Supporting information

Supplemental File 1: Detailed tables of demographics, baseline characteristics, and digital CBT study outcomes

## Data Availability

Data can be made available upon reasonable request to the corresponding authors (MR and TDV).

## ACKNOWLEDGEMENTS

This research was funded by grants from the Center for Aging + Brain Health Innovation (CABHI), Bell Canada (Bell Cause pour la cause), and the Research Center of the Institut Universitaire de Gériatrie de Montréal (CRIUGM). MR has been supported by the Fonds de Recherche du Québec – Nature et Technologies (FRQNT) and fellowships from Concordia University. KG has been funded by grad students awards from the CIHR and local university awards (Concordia University, Université de Montréal). We acknowledge the contributions of the following students who assisted in participants’ recruitment and data collection: Mila Goulon, Victoria D’Amours, Chelsea Nkori and Ilona Scellier-Dekens.

## DATA AVAILABILITY

Data can be made available upon reasonable request to the corresponding authors (MR and TDV).

