## Supplemental File 1: Detailed tables of demographics, baseline characteristics, and digital CBT study outcomes for "Feasibility and efficacy of a digital cognitive behavioural therapy program for insomnia and anxiety for older adults"

|  | **Group comparison**  **(whole sample)** | **Group comparison**  **(eCBT+ group only)** |
| --- | --- | --- |
| Age | *W* = 364, *p*= .50 | *W* = 137, *p*= .84 |
| Women | *χ^2^*(1, *N*= 80) = 0.28, *p*= 0.60 | *χ^2^*(1, *N*= 38) = 0.14, *p*= 0.71 |
| Education | *χ^2^*(2, *N*= 80) = 0.28, *p*= 0.87 | *χ^2^*(2, *N*= 38) = 4.25, *p*= 0.12 |
| Married/partnered | *χ^2^*(1, *N*= 80) = 0.22, *p*= 0.64 | *χ^2^*(1, *N*= 38) = 0.16, *p*= 0.69 |
| Taking sleep medication | *χ^2^*(1, *N*= 80) = 0.51, *p*= 0.47 | *χ^2^*(1, *N*= 38) = 0.68, *p*= 0.41 |
| Self-reported sleep apnoea | *χ^2^*(1, *N*= 80) < 0.01, *p*= 0.94 | *χ^2^*(1, *N*= 80) < 0.01, *p*= 1 |
| Technology usage | *χ^2^*(3, *N*= 80) = 0.89, *p*= 0.83 | *χ^2^*(3, *N*= 80) = 2.23, *p*= 0.53 |
| Sleep efficiency at baseline | *t*(8.32) = -0.37, *p*= .72 | *t*(10.04) = -0.23, *p*= .82 |
| Insomnia severity | *t*(10.46) = 0.21, *p*= .84 | *t*(12.77) = -0.20, *p*= .84 |
| Anxiety severity | *t*(11.25) = -1.03, *p*= .32 | *t*(15.74) = -1.47, *p*= .16 |

**Table S1.** Comparison of demographic and baseline characteristics between participants who dropped out and (1) the whole sample (left column) and (2) those who were randomized in the eCBT+ group (right column). Statistical tests (χ^2^ for categorical variables and *t*-test or Mann-Withney for continuous variables) were used to assess group differences (*p*< .05).

|  | eCBT+ group | | |  | Waitlist group | | |  | Time x Group | | | | |
| --- | --- | --- | --- | --- | --- | --- | --- | --- | --- | --- | --- | --- | --- |
| Outcome/Timepoint | *n* | Mean (SD) | Cohen’s d_within_ [95% CI] |  | *n* | Mean (SD) | Cohen’s d_within_  [95% CI] |  | *F* value | β | SE | *p* value | Cohen’s d_between_  [95% CI] |
| Sleep efficiency (%) |  |  |  |  |  |  |  |  |  |  |  |  |  |
| T0 | 34 | 69.78 (11.58) | 1.20 [0.67, 1.73] |  | 31 | 67.34 (12.90) | 0.17 [-0.21, 0.55] |  | *F*(1, 51.48) =24.37 | -10.66 | 2.16 | *p*< .001 | 1.33 [0.31, 2.35] |
| T1 | 24 | 81.33 (6.92) |  |  | 27 | 69.45 (13.41) |  |  |  |  |  |  |  |
| Insomnia severity |  |  |  |  |  |  |  |  |  |  |  |  |  |
| T0 | 35 | 16.09 (4.31) | 0.96 [0.48, 1.43] |  | 31 | 16.58 (5.27) | 0.04 [-0.33, 0.40] |  | *F*(1, 55.15) =15.41 | 5.06 | 1.29 | *p*< .001 | 0.92 [0.31, 1.52] |
| T1 | 25 | 10.56 (4.01) |  |  | 29 | 16.69 (5.33) |  |  |  |  |  |  |  |
| Anxiety severity |  |  |  |  |  |  |  |  |  |  |  |  |  |
| T0 | 35 | 9.66 (5.13) | 0.76 [0.31, 1.21] |  | 31 | 10.23 (5.51) | 0.11 [-0.26, 0.48] |  | *F*(1, 55.11) =10.45 | 3.43 | 1.06 | *p*< .01 | 0.93 [0.10, 1.77] |
| T1 | 25 | 5.40 (4.37) |  |  | 28 | 10.25 (5.53) |  |  |  |  |  |  |  |

**Table S2.** Effects of the eCBT+ program on sleep-diary sleep efficiency, insomnia severity (ISI), and anxiety symptoms (GAI). Individuals self-reporting sleep apnoea were excluded from both the eCBT+ and waitlist groups. Values are presented as mean with standard deviation (SD) for each group and timepoint. Linear model estimates (*F* value, β and standard error (SE)) and associated *p*-values are reported with effect sizes (Cohen’s *d*) for each outcome. A positive Cohen’s *d* indicates symptoms improvement, whereas a negative effect size indicates symptom worsening.

|  |  | **Pre-treatment** |  | **Post-treatment** |  | **Time** | | | |
| --- | --- | --- | --- | --- | --- | --- | --- | --- | --- |
| Outcome | *n* | Mean (SD) |  | Mean (SD) |  | β | SE | *p* value | Cohen’s d_within_ [95% CI] |
| Sleep efficiency (%) | 36 | 68.66 (12.38) |  | 78.14 (12.17) |  | -9.18 | 1.49 | *p<* .001 | 1.41 [0.91, 1.91] |
| Insomnia severity (ISI) | 37 | 15.97 (3.82) |  | 11.30 (4.90) |  | 5.02 | 0.86 | *p<* .001 | 1.25 [0.78, 1.72] |
| Anxiety severity (GAI) | 37 | 10.08 (5.16) |  | 6.86 (5.01) |  | 3.16 | 8.38 | *p<* .001 | 1.03 [0.56, 1.51] |

**Table S3.** Descriptive statistics and effect estimates for participants receiving the eCBT+ intervention. T2 data from participants assigned to the eCBT+ group were combined with T3 data from participants initially on the waitlist who subsequently received the eCBT+ program. Mean ± standard deviation (SD) are shown for pre- and post- treatment time points. Linear model estimates (β and standard error (SE)) and associated *p*-values are reported with within group effect sizes (Cohen’s *d*) for each outcome. A positive Cohen’s *d* indicates symptoms improvement, whereas a negative effect size indicates symptom worsening.

| **Author** | **Year** | **Total sample size (% Female)** | **Mean age, years (SD)** | **Other**  **health conditions** | **Study design** | **N^o^**  **of sessions** | **Retention rate (%)** | **Change in sleep efficiency post-intervention (%)** | **Change in ISI score post-intervention** | **Change in anxiety score post-intervention** |
| --- | --- | --- | --- | --- | --- | --- | --- | --- | --- | --- |
| Batterham et al.[1] | 2017 | 1149 (73.6) | 42.73 (12.21) | Depression | 2 arms (SHUTi, active control) | 6 | 43.2 | / | -8.57, *d* = 1.10 | GAD-7: -2.62, *d* = 0.50 |
| Brückner et al. [2] | 2025 | 74 (85.1) | 43.8 (12.0) | Shift work sleep disorders | 2 arms (SleepCare, digital psychoeducational self-help guidebook for shift workers) | 6 | 86 | / | -7.8, *d* = 1.78 | GDS-15: -3.4 |
| Chan et al. [3] | 2023 | 320 (73) | 27.27 (7.24) | Depression | 2 arms (proACT-S, waitlist) | 6 | 65.3 | / | -1.58 | HADS-A: n.s |
| Chan_et_al. [4] | 2024 | 129 (76) | 34.09 (12.05) | None | 5 arms (Sleep Sensei-therapist, Sleep Sensei -assitant, Sleep Sensei -chatbot, Sleep Sensei -unguided, sleep hygiene) | 6 | 66.7 | / | -6.12 | GAD-7: n.s. |
| Cheng et al. [5] | 2019 | 658 (79) | 45.1 (15.45) | Depression | 2 arms (Sleepio, online sleep hygiene education) | 6 | 68.3 | / | -10 | / |
| Christensen et al. [6] | 2016 | 1149 | 42.73 (12.21) | Depression | 2 arms (SHUTi, active control) | 6 | 43.2 | / | -8.6, *d* = 1.10 | GAD-7: -2.6, *d* = 0.5 |
| Clara et al. [7] | 2025 | 154 (94.8) | 47.0 (9.5) | Cancer | 2 arms (OncoSleep, waitlist) | 6 | 90.9 | 20.1, *d* = 1.35 | -11, *d* = 2.56 | HADS-A: -2.6, *d* = 0.77 |
| Ell et al. [8] | 2024 | 46 (80.4) | 39.7 (12.1) | Shift work sleep disorders | 2 arms (SleepCare, waitlist) | 6 | 82.6 | 5.1, *d* = 0.74 | -5.4, *d* = 1.25 | STAI-state, -3.5: *d* = 0.69  STAI-trait, -2.1: *d* = 0.47 |
| Espie et al. [9] | 2012 | 164 (73.2) | 49.0 (13.5) | None | 3 arms (Sleepio, active control, TAU) | 6 | 85.5 | 19.5, *d* = 1.28 | / | / |
| Felder et al. [10] | 2020 | 208 (100) | 33.6 (3.7) | Pregnant | 2 arms (Sleepio, TAU) | 6 | 93.3 | 8.4 | -5.9, *d* = 1.03 | GAD-7: -1.9, *d* = 0.42 |
| Glozier et al. [11] | 2019 | 87 (0) | 58.35 (6.2) | Depression | 2 arms (SHUTi, online psychoeducation) | 6 | 88.9 | / | -7.3, *d* = 0.62 | STAI-trait: n.s. |
| Hagatun et al. [12] | 2019 | 181 | 44.9 (13.0) | None | 2 arms (SHUTi, active control) | 6 | 81.1 | 15.15, d = 0.78 | -8.67, *d* = 1,77 | / |
| **Author** | **Year** | **Total sample size (% Female)** | **Mean age, years (SD)** | **Other**  **health conditions** | **Study design** | **N^o^**  **of sessions** | **Retention rate (%)** | **Change in sleep efficiency post-intervention (%)** | **Change in ISI score post-intervention** | **Change in anxiety score post-intervention** |
| Hinterberger et al. [13] | 2024 | 57 (68) | 45.67 (16.38) | None | 2 arms (NUKKUAA, waitslit) | 6 | 90 | n.s. | -5, *d* = 1.23 | / |
| Horsch et al. [14] | 2017 | 151 (62.3) | 39.66 (13.44) | None | 2 arms (Sleepcare, waitlist) | 6 | 61 | 7.2, *d* = 0.71 | -6.5, *d* = 0.66 | / |
| Kallestad et al. [15] | 2021 | 101 (75) | 41.35 (11.5) | None | 2 arms (SHUTi, face to face CBTi) | 6 | 89.8 | n.s. | -5.7, *d* = 1.2 | / |
| Kalmbach et al. [16] | 2020 | 91 (100) | 29.03 (4.16) | Pregnant | 2 arms (Sleepio, online sleep hygiene education) | 6 | 93.5 | / | -4.91 | / |
| Kyle et al. [17] | 2020 | 410 (86.6) | 52.45 (11.45) | None | 2 arms (Sleepio, waitlist) | 6 | 82.9 | / | -7.75, *d* = 1.57 | GAD-2: -1.01, *d* = 0.39 |
| Lee et al. [18] | 2025 | 106 (67.9) | 35.5 | None | 2 arms (WELT-I, online sleep hygiene education) | 6 | 91.7 | 12.53 | -5.33 | GAD-7: -1.15 |
| Lorenz et al. [19] | 2019 | 56 (70) | 42.88 (18.68) | None | 2 arms (dCBTi, waitlist) | 6 | 86.2 | 15.09 | -7.58, *d* = 1.79 | BSI-Anxiety: n.s. |
| Mason et al. [20] | 2023 | 120 (75) | 46.49 (13.13) | Anxiety | 2 arms (CBT for insomnia and CBT for anxiety) | 4 | 93.2 | 14.32, *d* = 0.36 | -11.9, *d* = 0.72 | GAD-7: -8.19, *d* = 0.29 |
| Maurer et al. [21] | 2025 | 56 (85.7) | 45.55 (13.70) | None | 2 arms (Somnio, Malio - a digital sleep monitoring application) | 10 | 93.1 | 7.38 | -6.04, *d* = 1.08 | HADS-A: n.s. |
| McGrath et al. [22] | 2017 | 134 (61.2) | 59 (10.9) | Blood pressure 130-160/<110mm Hg | 2 arms (Sleepio, vascular risk factor education) | 6 | 80.6 | 5.0 | -4.5 | BAI: -2.2 |
| Park et al. [23] | 2025 | 54 (72.2) | 38.75 (12.4) | None | 2 arms (WELT-I, online sleep hygiene education) | 6 | 84.8 | 14.4, *d* = 0.68 | n.s. | GAD-7: n.s. |
| Ritterband et al. [24] | 2009 | 44 (77.3) | 44.86 (11.03) | None | 2 arms (SHUTi, waitlist) | 6 | 95.5 | 16 | -9.14 | / |
| Ritterband et al. [25] | 2012 | 28 (85.7) | 56.7 (11.7) | Cancer | 2 arms (SHUTi, waitlist) | 6 | 100 | 13.51, *d* = 0.72 | -8.9, *d* = 1.85 | HADS-A: n.s. |
| **Author** | **Year** | **Total sample size (% Female)** | **Mean age, years (SD)** | **Other**  **health conditions** | **Study design** | **N^o^**  **of sessions** | **Retention rate (%)** | **Change in sleep efficiency post-intervention (%)** | **Change in ISI score post-intervention** | **Change in anxiety score post-intervention** |
| Ritterband et al. [26] | 2017 | 303 (71.9) | 43.28 (11.59) | None | 2 arms (SHUTi, online patient education) | 6 | 88.1 | 12.35, *d* = 0.92 | *d* = 1.90 | / |
| Ritterband et al. [27] | 2025 | 311 (68.5) | 66.3 (7.2) | None | 3 arms (SHUTi OASIS, SHUTi OASIS and stepped support, online sleep hygiene education) | 6 | 95 | 12.12, *d* = 1.19 | -5.57, *d* = 1.04 | / |
| Sato et al. [28] | 2022 | 270 (41.5) | 50.43 (10.83) | None | 3 arms (dCBTi, 3 good things exercise, waitlist) | 4 | 77.5 | 5.85 | / | GAD-7: n.s. |
| Schuffelen et al. [29] | 2023 | 238 (67.6) | 43.73 (13.90) | None | 2 arms (Somnio, waitlist) | 10 | 89.8 | 17.34 | -9.07, *d* = 2.08 | STAI-trait: -5.5, d = 0.56 |
| Schuffelen et al. [30] | 2025 | 140 (85.7) | 39.76 (11.69) | Depression | 2 arms (Somnio and TAU, waitlist and TAU) | 10 | 77.1 | *d* = 0.63 | *d* = 1.94 | / |
| Schuffelen et al. [31] | 2025 | 207 (182) | 51.96 (12.97) | Chronic pain | 2 arms (Somnio and TAU, waitlist and TAU) | 10 | 88.3 | / | *d* = 1.18 | STAI-trait: *d* = 0.27 |
| Shin et al. [32] | 2024 | 98 (61.2) | 42.3 (12.95) | None | 2 arms (Somzz, sleep hygiene education) | 6 | 87.8 | 12.6 | -3.08 | GAD-7: -1.7 |
| Specht et al. [33] | 2025 | 290 (73.8) | 49.83 (14.12) | None | 2 arms (Somnovia and TAU, waitlist and TAU) | 6 | 81.9 | / | -6.7, *d* = 0.71 | GAD-7: -2.8, *d* = 0.56 |
| Sweetman et al. [34] | 2024 | 62 (82.2) | 52.46 (16.26) | None | 2 arms (Somnovia, online sleep hygiene education) | 5 | 90.3 | / | -9.21, *d* = 1.64 | GAD-7: -2.94, *d* = 0.5 |
| Vedaa et al. [35] | 2020 | 1721 (67.9) | 44.5 (13.8) | None | 2 arms (SHUTi, online sleep hygiene education) | 6 | 67.3 | 11.7, d = 0.54 | -8.8, *d* = 1.21 | / |
| Watanabe et al. [36] | 2023 | 175 (58.3) | 44.15 (13.35) | None | 2 arms (dCBTi, waitlist) |  | 100 | 10.4 | / | / |
| Zhou et al. [37] | 2022 | 333 (100) | 59.5 (8.0) | None | 3 arms (SHUTi, SHUTi for black women, online sleep hygiene education) | 6 | 78.7 | 12.2 | -8.3 | / |

**Table S4.** Summary of randomized controlled trials reporting significant effects of digital cognitive behavioural therapy for insomnia on sleep efficiency, Insomnia Severity Index (ISI) or anxiety symptoms. For each study, demographic characteristics, retention rates, pre-to-post changes and between-group effect sizes (Cohen’s *d*) are presented. A positive Cohen’s *d* indicates symptoms improvement, whereas a negative effect size indicates symptom worsening. The table is adapted from Hwang et al. [38], incorporating additional data and newly published article from March 31, 2024 to October 14, 2025.

*N.B: BAI = Beck Anxiety Inventory, BSI-Anxiety = Brief Symptom Inventory – Anxiety subscale, GAD-7 = Generalized Anxiety Disorder 7 items, GDS = Geriatric Depression Scale, HADS-A = Hospital Anxiety and Depression Scale – Anxiety subscale, n.s. = non significant, STAI = State-Trait Anxiety Inventory, TAU = Treatment As Usual*
